# Changes in the number of public health nurses employed in local governments in Japan during the Covid-19 pandemic: A cross-sectional study

**DOI:** 10.1101/2022.02.06.22270346

**Authors:** Kazuya Taira, Rikuya Hosokawa, Misa Shiomi

## Abstract

**Objectives:** This study aims to clarify the recruitment of public health nurses in local governments in Japan during the Covid-19 pandemic.

**Study design:** A cross-sectional study.

**Methods:** A cross-sectional study of 150 local governments that have public health centers in Japan was conducted. The survey period was November to December 2021. The survey items were the number of full-time and part-time public health nurses (PHNs), the number of PHNs who resigned or retired from the job, and the number of PHN recruitment examinations for each year from 2017 to 2021. For all variables, the mean, standard deviation, maximum, and minimum values for each type of municipality and year were calculated, and a one-way analysis of variance was performed.

**Results:** The recovery rate was 54.0% (81/150). Although a statistically significant difference was not recorded in the change in employment status of PHNs from 2019 to 2020, during the year that COVID-19 infection began in Japan, the number of full-time PHNs increased by only 2.6 at the maximum, while the number of part-time PHNs was 5.2±8.3 to 10.8±9.6 (p = 0.61) for prefectures, from 13.6±13.1 to 21.5±34.8 (p = 0.23) for city, and from 16.8±26.8 to 52.3±132.5 (p = 0.70) for ward.

**Conclusions:** This study reveals that support for the increased workload due to COVID-19 is heavily dependent on part-time PHNs. Drastic change to the ideal way of the original countermeasure to Covid-19 in Japan or the supply of stronger human support to the public health center might be desired.

## Introduction

Since January 2020, a series of nationwide outbreaks of Covid-19 has occurred in Japan.^1^ In Japan, public health nurses working in public health centers owned by local governments were responsible for active epidemiological research, coordination of the polymerase chain reaction (PCR) testing, the control of admission to and discharge from hospitals, and the health management of patients recuperating at home.^2,3^ The workload increased with the number of patients, but the changes in the number of public health nurses was not researched. This study aims to examine the recruitment of public health nurses in local governments in Japan during the Covid-19 pandemic.

## Methods

A cross-sectional study was conducted of 150 local governments that have public health centers in Japan: 47 prefectures, 80 cities, and 23 special wards of Tokyo. The survey was carried out from November to December 2021, and it was executed by mail to the human resources department of each local government. The subjects were the number of full-time and part-time PHNs, the number of PHNs who resigned or retired from the job, and the number of PHN recruitment examinations for each year from 2017 to 2021. For all variables, the mean, standard deviation, maximum, and minimum values for each type of municipality and year were calculated, and a one-way analysis of variance was performed.

## Results

The recovery rate was 54.0% (81/150). Focusing on the results from 2020, when the COVID-19 outbreak began, the change in the number of full-time PHNs was from 112.3 ± 52.8 in 2019 to 112.6 ± 52.5 in 2020 (p = 0.99) for prefectures, from 106 ± 90.6 in 2019 to 107.6 ± 91.6 in 2020 (p = 0.99) for cities, and from 62.8 ± 23.5 in 2019 to 65.4 ± 25.3 in 2020 (p = 0.94) for wards.(Table 1) In addition, the change in the number of part-time PHNs was from 5.2±8.3 in 2019 to 10.8±9.6 in 2020(p = 0.61) for prefectures, from 13.6±13.1 in 2019 to 21.5±34.8 in 2020 (p = 0.23) for cities, and from 16.8±26.8 in 2019 to 52.3±132.5 in 2020 (p = 0.70) for wards. At the 5% level, there was a statistically significant difference in the number of PHNs who resigned or retired from work in all of the prefectures (p=0.00), cities (p=0.00), and wards (p=0.02). No significant differences were found in the number of recruitment examinations.

**Table 1.**
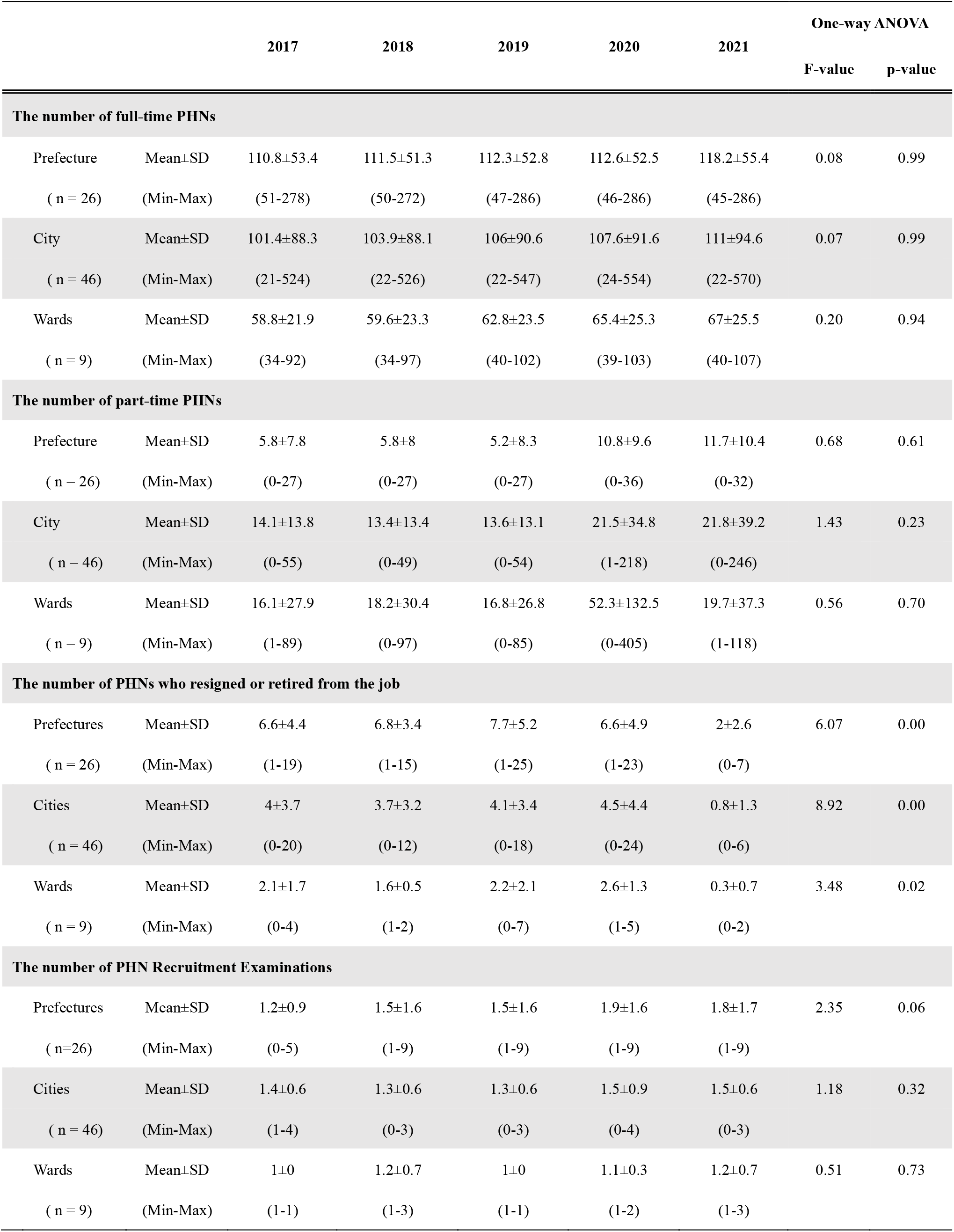
Changes in the employment status of public health nurses by type of local governments from 2017 to 2021

|  |  | 2017 | 2018 | 2019 | 2020 | 2021 | One-way ANOVA |  |
| --- | --- | --- | --- | --- | --- | --- | --- | --- |
|  |  |  |  |  |  |  | F-value | p-value |
| **The number of full-time PHNs** |  |  |  |  |  |  |  |  |
| Prefecture | Mean±SD | 110.8±53.4 | 111.5±51.3 | 112.3±52.8 | 112.6±52.5 | 118.2±55.4 | 0.08 | 0.99 |
| ( n = 26) | (Min-Max) | (51-278) | (50-272) | (47-286) | (46-286) | (45-286) |  |  |
| City | Mean±SD | 101.4±88.3 | 103.9±88.1 | 106±90.6 | 107.6±91.6 | 111±94.6 | 0.07 | 0.99 |
| ( n = 46) | (Min-Max) | (21-524) | (22-526) | (22-547) | (24-554) | (22-570) |  |  |
| Wards | Mean±SD | 58.8±21.9 | 59.6±23.3 | 62.8±23.5 | 65.4±25.3 | 67±25.5 | 0.20 | 0.94 |
| ( n = 9) | (Min-Max) | (34-92) | (34-97) | (40-102) | (39-103) | (40-107) |  |  |
| **The number of part-time PHNs** |  |  |  |  |  |  |  |  |
| Prefecture | Mean±SD | 5.8±7.8 | 5.8±8 | 5.2±8.3 | 10.8±9.6 | 11.7±10.4 | 0.68 | 0.61 |
| ( n = 26) | (Min-Max) | (0-27) | (0-27) | (0-27) | (0-36) | (0-32) |  |  |
| City | Mean±SD | 14.1±13.8 | 13.4±13.4 | 13.6±13.1 | 21.5±34.8 | 21.8±39.2 | 1.43 | 0.23 |
| ( n = 46) | (Min-Max) | (0-55) | (0-49) | (0-54) | (1-218) | (0-246) |  |  |
| Wards | Mean±SD | 16.1±27.9 | 18.2±30.4 | 16.8±26.8 | 52.3±132.5 | 19.7±37.3 | 0.56 | 0.70 |
| ( n = 9) | (Min-Max) | (1-89) | (0-97) | (0-85) | (0-405) | (1-118) |  |  |
| **The number of PHNs who resigned or retired from the job** |  |  |  |  |  |  |  |  |
| Prefectures | Mean±SD | 6.6±4.4 | 6.8±3.4 | 7.7±5.2 | 6.6±4.9 | 2±2.6 | 6.07 | 0.00 |
| ( n = 26) | (Min-Max) | (1-19) | (1-15) | (1-25) | (1-23) | (0-7) |  |  |
| Cities | Mean±SD | 4±3.7 | 3.7±3.2 | 4.1±3.4 | 4.5±4.4 | 0.8±1.3 | 8.92 | 0.00 |
| ( n = 46) | (Min-Max) | (0-20) | (0-12) | (0-18) | (0-24) | (0-6) |  |  |
| Wards | Mean±SD | 2.1±1.7 | 1.6±0.5 | 2.2±2.1 | 2.6±1.3 | 0.3±0.7 | 3.48 | 0.02 |
| ( n = 9) | (Min-Max) | (0-4) | (1-2) | (0-7) | (1-5) | (0-2) |  |  |
| **The number of PHN Recruitment Examinations** |  |  |  |  |  |  |  |  |
| Prefectures | Mean±SD | 1.2±0.9 | 1.5±1.6 | 1.5±1.6 | 1.9±1.6 | 1.8±1.7 | 2.35 | 0.06 |
| ( n=26) | (Min-Max) | (0-5) | (1-9) | (1-9) | (1-9) | (1-9) |  |  |
| Cities | Mean±SD | 1.4±0.6 | 1.3±0.6 | 1.3±0.6 | 1.5±0.9 | 1.5±0.6 | 1.18 | 0.32 |
| ( n = 46) | (Min-Max) | (1-4) | (0-3) | (0-3) | (0-4) | (0-3) |  |  |
| Wards | Mean±SD | 1±0 | 1.2±0.7 | 1±0 | 1.1±0.3 | 1.2±0.7 | 0.51 | 0.73 |
| ( n = 9) | (Min-Max) | (1-1) | (1-3) | (1-1) | (1-2) | (1-3) |  |  |

## Discussion

Although a statistically significant difference was not recorded in the change in employment status of PHNs, the number of full-time PHNs increased by only 2.6, even in the most local government type, while the number of part-time PHNs almost doubled. The number of recruitment examinations has not increased, and this study revealed that support for the increased workload due to COVID-19 is heavily dependent on part-time PHNs. Traditionally, most workers in Japan retire at the end of March. Hence, the number of retirees was statistically significant because a survey was conducted in the middle of the 2021 fiscal year. External support and the employment of part-time PHNs are among the measures to prevent dysfunction in public health centers;^4^ however, there are challenges such as maintaining a stable supply and difficulty in timely employment procedures against the speed of infection. As of January 2022, Japan is observing an unprecedented number of infections^1^ due to the highly infectious Omicron variant (B 1.1.529),^5^ and public health centers are on the verge of dysfunction. Although observation of Omicron has shown lower rates of disease severity and mortality,^6,7^ a drastic change to the original countermeasures to Covid-19 in Japan or the supply of stronger human support to public health centers might be desired.

## Data Availability

Participants of this study did not agree for their data to be shared publicly, so supporting data is not available.

## Acknowledgements

Ethics committee of the Kyoto University Graduate School and Faculty of Medicine, Kyoto University Hospital waived ethical approval for this work. This study was funded by the research support fund “Kusunoki 125” by the university to which the author belongs.

